# Effect of intermittent pringle manipulation on serum potassium concentration during laparoscopic hepatectomy: a self-controlled study protocol

**DOI:** 10.1101/2024.08.22.24312356

**Authors:** Yan Weng, Ziqi Shang, Yan Wang, Xiaojuan Liu, Yuling Tang, Hua Zhang, Chunmei Wu, Wenjie Mao, Qing Zhong

## Abstract

**Background:** Intermittent Pringle Manipulation (IPM) is among the most common methods used for controlling blood loss during hepatectomy. Ischaemia–reperfusion injury has also been associated with IPM. Ischaemic injury exposes the liver cells to hypoxia, adenosine triphosphate depletion, pH changes, and cellular metabolic stress, all of which can lead to cell damage and death. Reperfusion injury is caused by microcirculatory dysfunction, hypoxia, oxidative stress, and apoptosis. The pathophysiological mechanism of ischaemia-reperfusion injury is hyperkalaemia. Hyperkalaemia is closely related to the electrophysiological activity of the myocardium. Acute hyperkalaemia is associated with life-threatening ventricular arrhythmia and sudden cardiac arrest. Therefore, it is necessary to observe changes in patient serum potassium concentrations during IPM to provide a reference for developing a secure anaesthesia management approach.

**Methods and analysis:** This was a single-centre, open, non-interventional, self-controlled study. All eligible consecutive patients were recruited from a regional medical centre and scheduled for elective hepatectomy. There was no control group; all participants were continuously enrolled from 1 October 2023 to 31 May 2025. The primary outcome was the perioperative serum potassium concentration during IPM. Secondary outcomes included perioperative electrocardiogram changes, lactic acid status, postoperative serum potassium concentration, alanine amine transferase, and aspartate amine transferase peaks, adverse events, serious adverse events, and postoperative hospital stay. These parameters were statistically compared. Subgroup analysis will be performed according to liver disease type and duration of IPM.

**Discussion:** Our finding will provide a reference for developing a secure anaesthesia management approach for anesthesiologists.

**Ethics and dissemination:** The Biomedical Ethics Review Committee of the People’s Hospital of Jianyang City approved the study protocol (ethics reference: JY202383). All relevant ethical guidelines were followed in this study. The findings will be disseminated in peer-reviewed journals, publicly available reports to be published online, and academic conferences.

**Trial Registration:** This trial was registered on 25 August 2023 in the Chinese Clinical Trial Registry (http://www.chictr.org.cn), ChiCTR2300074753.

## Introduction

Hepatectomy is an important surgical procedure for the treatment of various liver diseases.[1, 2] The liver is an important reservoir of blood and receives approximately 25% of the resting cardiac output. The hepatic artery is responsible for 25–30% of the blood supply to the liver, whereas the portal vein is responsible for 70–75%, they each deliver 50% of the total oxygen to the liver.[3] Owing to the large amount of blood stored in the liver, controlling intraoperative bleeding is crucial in hepatectomy. Various methods have been used to control bleeding. Intermittent printing manipulation (IPM) is one of the most common bleeding control strategies.[4, 5]

IPM, which is a cycle of 15-min inflow occlusion of the portal vein and hepatic artery, followed by 5-min reperfusion, involves cutting off the blood supply to the liver to reduce bleeding during hepatectomy.[6] IPM is a crucial technique applied in hepatectomy and has been considered the gold standard method for controlling bleeding during hepatectomy.[7] It has been proven to not affect the long-term survival of patients with hepatocellular carcinoma.[8]

Normoxia-ischaemia-reperfusion occurs during IPM,[9] resulting in ischaemia-reperfusion injury (IRI) when blood flow is suddenly restored in the IPM cycle of 5-min reperfusion.

During the normoxic period, the liver performs several vital functions, including metabolism and detoxification. The ischaemic state induces anaerobic metabolism, leading to a lower level of ATP production and failure of ion-exchange channels such as sodium-potassium pumps (Na^+^-K^+^-ATPase pumps) and calcium pumps (Ca^2+^-ATPase pumps). The failure of ion exchange channels leads to cell swelling and impaired cytoplasmic enzymatic activity. During reperfusion, mitochondrial damage and an electrolyte imbalance in the reperfusion state promote oxidative stress. Simultaneously, the generation of reactive oxygen species (ROS) leads to cell death.[10] These pathological processes lead to liver acidosis, electrolyte disturbances, and increased serum K^+^ concentration ([K^+^]). Meanwhile, the large difference in [K^+^] inside and outside the cells[11] leads to the introduction of K^+^ into the plasma after cell necrosis, which also increases [K^+^].

K^+^ is an important ion in the electrophysiological activities of the myocardium.[12, 13] HK, defined as serum potassium concentration > 5.5 mEq /L,[14, 15] is closely related to electrocardiogram (ECG), including but not limited to peaked T waves, shortened QT interval, lengthening of PR interval, and QRS duration.[16-21] In the study by Wetmore JB et al,[15] nearly one in five patients with K ≥6.0 mEq/L, and nearly one in three patients with K ≥7.0 mEq/L, died, compared with only about one in eighty patients with a K of 3.5–5.0 mEq/L. Acute HK is associated with life-threatening ventricular arrhythmia and sudden cardiac arrest.[22] Postreperfusion hyperkalaemia is associated with several intraoperative and postoperative complications, including post-reperfusion syndrome, cardiac arrest,[23] intraoperative death, and postoperative mortality.[24-26] The incidence of intraoperative cardiac arrest during adult liver transplantation is 1.0%,[27] 2.1%,[28] or 5.5%[29] and the main aetiology of intraoperative cardiac arrest is hyperkalaemia.[23, 27]

Although there are no neohepatic stages during hepatectomy, IRI remains an important problem which can lead to HK. In 1972, Hall et al. conducted a study on HK after a temporary blockade of the portal vein and hepatic artery opening.[30] They concluded that temporary occlusion of the hepatic artery and portal vein in dogs causes transient but severe HK, which can cause cardiotoxicity even when the duration of circulatory arrest is only 10 min. Clinical interventional studies are ethically limited because participants with high serum potassium concentrations are unsafe.[20, 21, 31]

However, with the development of surgical treatments, subsequent studies have mainly focused on the safety of IPM during hepatectomy, including intraoperative blood transfusion, postoperative liver function impairment, postoperative complications, and postoperative liver tumor recurrence,[32-34] and the HK is gradually being neglected.

Based on the theoretical foundation of real-world research, there was no intervention and there was no violation of the ethical principles of medical research. The study protocol was reasonable and the results of this trial may be clinically significant.

## Methods and aims of the study

### Study design

This was a single-centre, open, non-interventional, self-controlled study. Serum potassium concentration was monitored before IPM and within 5 min after the vessels were opened to evaluate the influence of IPM on serum potassium concentration. All participants were continuously enrolled from 1 October 2023 at the Hospital of Jianyang. This study was conducted in accordance with the principles of clinical research. The results are reported in accordance with the SPIRIT checklist[35]. The first participant was enrolled on 9 November 2023, and we got a written informed consent from him. We plan to conclude the follow-up of the last patient on 31 May 2025.

### Study setting

The study was conducted at the People’s Hospital of Jianyang City, Chengdu, Sichuan, China. The People’s Hospital of Jianyang is a public tertiary Grade A hospital that performs more than 500 hepatectomies annually. The Department of Anaesthesiology has a strong scientific research capacity and nine anaesthesiologists participated in the study. Investigators in the trial are responsible for completing the operation, managing and interpreting the data, writing the final report, and making the decision to submit the report for publication and have ultimate authority over any of these activities.

### Objectives

Primary objective:

The primary objective is to observe the effect of intermittent pringle manipulation on serum potassium concentration during laparoscopic hepatectomy.

Secondary objective:

The secondary objective is to observe the security of IPM.

### Recruitment

This recruitment will be conducted from 1 October 2023 to 31 May 2025. There was no control group; therefore, the number of enrolled patients was not limited. All patients who met the inclusion criteria were enrolled. The day before surgery, along with the pre-anaesthetic assessment anaesthesiologist, a trained research assistant will inform the patient about the purpose, content, and points for attention of the trial; voluntariness, benefits, and risks related to the study; principles of data protection and handling; and dissemination plans for the trial results. The patient and/or immediate family may express their willingness to participate and sign a written informed consent form. Figure 1 presents the study procedure and schedule for the outcome assessment.

Based on the anatomical structure, venous drainage of the liver occurs through the hepatic veins directly into the inferior vena cava (IVC) and IVC drainage into the right atrium (RA). According to Kin et al.,[23] serum potassium levels peaked 1 min after reperfusion in liver transplantation. Therefore, we collected mixed venous blood samples from the RA within 1 min of opening the vessel during IPM. A central venous catheter (CVC) was placed in the RA under ultrasound guidance for every enrolled patient to collect mixed venous blood from the RA.

The maximum duration of normothermic ischaemia during liver surgery, which is classically set at 60 min, can be safely extended to 120 min by using intermittent pedicular clamping.[36] Based on the results of Belghiti et al.,[37] the use of ischaemia cycles not exceeding 15 min in IPM was based on the fact that this period was the optimal ischaemic period to avoid irreversible liver cell damage, as demonstrated in experimental models.[38]

### Inclusion criteria

The inclusion criteria were as follows: 1) patients aged ≥18 and ≤80 years, 2) American Society of Anaesthesiologists classification I to III, 3) elective hepatectomy was scheduled, 4) operative time ≤5 h, 4) 3.5 mmol/L≤K^+^≤5.5 mmol/L before the operation.

### Exclusion criteria

The exclusion criteria were as follows: 1) refusal to participate in the study, 2) requirement of CPR or electrical defibrillation due to special conditions during the operation, 3) patients with electrolyte disturbances before surgery

### Study procedure

Anesthesia management

All patients were administered general anaesthesia with tracheal intubation. Intraoperative monitoring included noninvasive measurement of blood pressure, electrocardiography, pulse oxygen saturation, radial artery blood pressure, and nasopharyngeal temperature. Intravenous anaesthesia was induced using midazolam, sufentanil, etomidate, and rocuronium. After successful tracheal intubation, the patient received mechanical ventilation and end-tidal carbon dioxide was maintained between 35 and 45 mmHg. Anaesthesia was maintained using propofol, remifentanil, and cisatracurium to maintain a bispectral index value of 40–60 (index ranges from 1 to 100; higher values indicate shallower depths of sedation). Hypotension (mean arterial pressure <65 mmHg or a 20% decrease from baseline) and bradycardia (heart rate <50 beats/min) were induced. A thermal blanket was used during the operation to maintain the nasopharyngeal temperature above 36 °C.

### Data collection

An outcome assessor collected data from the electronic medical record system. All participants were scheduled for selective hepatectomy under general anaesthesia. Our planned recruitment period was from 1 October 2023 to 31 May 2025 with a postoperative follow-up of 7 days.

Demographic data:

1. Age, year
2. Sex, male/female
3. Body mass index, kg/m^2^
4. Comorbid conditions
5. ASA physical status classification
6. Operation duration
7. Anaesthesia duration
8. Postoperative outcomes (post-anaesthesia care unit (PACU)/intensive care unit (ICU))
9. Intraoperative blood loss
10. Preoperative liver function indications: ALT and its peak, AST and its peak
11. Preoperative cirrhosis
12. Preoperative hepatitis B infection
13. Perioperative blood transfusion or not

Preoperative laboratory tests □

Serum potassium concentration

We used the machine SIMENS ADVIA 2400 machine with a sensitivity of 1 mmol/L to monitor kalemia.

Perioperative data □

1. Basal serum potassium concentration (T_0_): Data were obtained from blood gas analysis and blood was obtained from the central venous catheter, which was placed at the junction of the inferior vena cava and right atrium.
2. Serum potassium concentration at the end of the 1^st^ IPM (T_1_): Data were recorded as T_1_. Blood was obtained 5 s after vessel opening.
3. Serum potassium concentration at the end of the 2^nd^ IPM (T_2_): The data were recorded as T_2_.
4. Serum potassium concentration at the end of the 3^rd^ IPM (T_3_): Data were recorded as T_3_.
5. ……
6. Electrocardiogram changes during operation
7. Lactic acid status
8. Use of potassium-containing drugs and infusion

Postoperative data:

1. Serum potassium concentration
2. ALT, AST, and their peaks
3. Adverse events (AE) and serious adverse events (SAE)
4. Postoperative hospital stay (day)

### Outcome measures

Data management

Trained outcome assessors recorded data in a case report form (CRF) in a timely, complete, and accurate manner. The CRF forms were checked by outcome assessors. They then entered the data into Excel software. Statistical analyses were performed after the trial. The Biomedical Ethics Committee of the People’s Hospital of Jianyang was responsible for monitoring the safety and procedures of this study.

### Statistical analysis

This trial was designed to observe the effects of IPM on serum potassium concentrations. Changes in serum potassium levels in the enrolled patients were summarised in a self-controlled study.

For the primary outcome, serum potassium concentration and repeated measures ANOVA will be used to analyse the changes in serum potassium at T0, T1, T2, and T3.

For demographic data, continuous variables will be presented as the mean ± standard deviation or median with its range based on normality testing and compared using an independent sample Student’s T or Mann–Whitney U test. Categorical variables will be presented as numbers (percentages) and were compared using the chi-square or Fisher’s exact test.

The significance of postoperative categorical complications containing AE and SAEs will be analysed using the chi-square or Fisher’s exact test or will be tabulated and summarised using descriptive statistics.

A p-value of <□0.05 will be taken as the threshold for statistical significance.

### Oversight and monitoring

Anaesthesiologists and statisticians will be responsible for overseeing and monitoring the entire trial. Subject diagnosis, recruitment, and informed consent will be handled by assistant investigators. The progress, relevant events, and data quality of the trial will be evaluated by the Biomedical Ethics Review Committee of People’s Hospital of Jianyang. There will be no interim analysis. If individual participants reported SAEs during the trial, the principal investigator decides would terminate the trial. The study team will report two monthly data points to the committee to monitor the progress and quality of the study.

### Patient and public involvement

All the research questions and outcome measures were based on objective test results and facts. Patients and the public will not be involved in the study design, recruitment, implementation, reporting, or assessment. The results of this study will be disseminated to the public through academic papers and conferences.

## Data Availability

No datasets were generated or analysed during the current study. All relevant data from this study will be made available upon study completion.

## Discussion

To reduce bleeding, IPM plays an important role in laparoscopic hepatectomy. In clinical practice, ECG changes are always observed within 10 min after vascular opening in patients using an IPM during hepatectomy. In some serious cases, patients may experience ventricular fibrillation or cardiac arrest. As reported by Shang Z et al[39], the rapid change of serum potassium level is an important cause of ventricular fibrillation and cardiac arrest in liver resection surgery when IPM is used to control bleeding.

There are three main causes account for HK: (1) IRI causes the release of congested blood, which can cause marked acidaemia, mobilising potassium from a large intracellular pool. (2) The liver releases large amounts of intracellular potassium under stress. (3) Endogenous metabolites that accumulate in the liver graft during ischaemia are returned to the body. HK can have fatal consequences, such as severe arrhythmias or asystole. Therefore, vigilant monitoring of blood potassium levels during IPM is important to provide a reference for developing safe anaesthesia management procedures.

We must acknowledge the Strengths and limitations of this study. (1) This was an observational study with no intervention, and there was no harm to patients. (2) Based on real-world research methods, the results are close to actual clinical situations. (3) The use of visualisation technology to locate the central venous catheter can provide more reliable research results. (4). To avoid the harm of multiple blood collections to patients, a blood test before vessel opening during IPM was not performed, which will not provide a more comparable self-controlled result.

## Status and timeline of the study

This study began to recruit patients on 1 October 2023. The follow-up of all patients will be completed on 31 May 2025. Analysis is expected to start during the fall of 2025 after all data being collected. The study is set to be finished by the winter of 2025.

## Ethics and dissemination

The Biomedical Ethics Review Committee of the People’s Hospital of Jianyang City approved the study protocol (ethics reference: JY202383). All relevant ethical guidelines were followed in this study. The findings will be disseminated in peer-reviewed journals, publicly available reports to be published online, and academic conferences.

## Author contributions Statement

**Conceptualization:** Ziqi Shang and Yan Wang.

**Funding acquisition:** Yan Weng.

**Methodology:** Xiaojuan Liu, Hua Zhang, Chunmei Wu, and Yuling Tang.

**Project administration**: Wenjie Mao.

**Resources:** Yan Weng.

**Supervision:** Yan Weng

**Writing-original draft:** Yan Weng, Ziqi Shang and Yan Wang.

**Writing-review & editing:** Wenjie Mao and Qing Zhong.

All authors contributed to the manuscript, read it, and agreed to its publication.

Figure Legend:

Figure 2 – Flow chart

